# How well it works: Benchmarking performance of GPT models on medical natural language processing tasks

**DOI:** 10.1101/2024.06.10.24308699

**Authors:** Kathryn Rough, Hui Feng, Paul B Milligan, Francesco Tombini, Tom Kwon, Khaldoun Zine El Abidine, Christina D Mack, Benjamin Hughes

## Abstract

**Importance:** The ability of large language models (LLMs) to generate high-quality, human-like text has been accompanied with speculation about their application in healthcare, alongside ethical and safety concerns.

**Objective:** Evaluate LLM performance on medical natural language processing (NLP) tasks, benchmarked against other commercially available tools.

**Design:** Observational study to evaluate and compare model performance. All models were commercially available and were evaluated without modification.

**Setting:** The Text Analysis Coding (TAC) 2017 challenge was used to assess ability to perform medical coding using standard MedDRA preferred terms. Text from 55 publicly available de-identified medical transcription reports were annotated to identify pre-defined medical concepts (age, disease/symptom, body structure, medication name, and medication dosage).

**Participants:** Publicly available, de-identified adverse event and medical transcription reports were used for evaluation.

**Exposures:** For each task, general LLMs (GPT-3.5-turbo, GPT-4) were compared to commercially available healthcare NLP tools (Microsoft Text Analytics for Health, Amazon Comprehend Medical, IQVIA API Marketplace).

**Main Outcomes and Measures:** For each NLP task, sensitivity, positive predictive value (PPV) and F1 score were calculated. Because GPT models had variable outputs, the range of metrics over 5 trials is reported.

**Results:** For MedDRA coding, GPT-4 had similar F1 score performance to healthcare NLP algorithms (GPT-4: 0.67 to 0.73; Microsoft Text Analytics for Health: 0.66, IQVIA API Marketplace: 0.72), while GPT-3.5-turbo had considerably lower performance (0.50 to 0.51). For medical information extraction, LLM performance varied widely across differing medical concepts; the highest F1 scores were for age (GPT-3.5-turbo: 0.82 to 0.83, GPT-4: 0.84 to 0.87) and medication name (GPT-3.5-turbo: 0.55 to 0.59, GPT-4: 0.70 to 0.76), while F1 scores for disease/symptom, body structure, and medication dosage were lower than those observed for the healthcare NLP tools. GPT-3.5-turbo and GPT-4 generally had lower sensitivity than comparators.

**Conclusions and Relevance:** In the absence of domain-specific fine tuning, GPT-4 performed similarly to healthcare-specific NLP tools on some tasks and less accurately on others; GPT-3.5-turbo was consistently less accurate than comparators. To maximize benefit and reduce risk of harm, robust quantitative evaluation for specific tasks should be performed prior to implementing LLMs in medical contexts.

## Introduction

There has been rapid progress in creating powerful large language models (LLMs) that are capable of answering questions, translating text, creating summaries, and more.^1,2^ These models have generated considerable excitement and speculation about potential healthcare applications, including support of clinical decision-making,^3^ preparation of manuscripts,^4^ and aiding clerical documentation activities.^5^ Alongside this potential, there are serious concerns about ethics of LLM use,^6^ generation of false or harmful information,^4^ and exacerbation of existing health inequalities.^4,7^

Despite enthusiasm for leveraging these tools and calls for evaluations in healthcare contexts,^8^ there are limited quantitative assessments of their utility for practical use cases. Emerging evidence shows that for several biomedical natural language processing (NLP) tasks, performance of general purpose LLMs can range from good to extremely poor.^9,10^

To facilitate an evidence-based assessment of appropriate use of these technologies in practice, the objective of this study is to evaluate the performance of a selection of commercially available LLMs against existing alternatives for medical data extraction and coding tasks.

## Methods

This observational study compared model performance of two general LLMs, GPT-3.5-turbo and GPT-4, with three specialized healthcare NLP tools: Microsoft Text Analytics for Health, Amazon Comprehend Medical, and IQVIA API Marketplace. All algorithms were commercially available; no new models were trained. These algorithms were benchmarked on two NLP tasks frequently performed in clinical research: MedDRA coding and medical entity extraction.

To assess algorithm ability to perform medical coding and harmonization, we used Task 4 of the publicly available 2017 Text Analysis Coding (TAC) challenge.^11^ Each comparator used to map free text descriptors of adverse drug reactions into the MedDRA® (Medical Dictionary for Regulatory Activities) preferred term ontology, which is commonly used in pharmacovigilance and medications safety surveillance.

To assess algorithm ability to extract categories of medical information from free text (i.e., medical entity recognition), we created a benchmarking dataset using publicly available de-identified medical transcription reports. A sample of 55 reports were annotated by trained nurses. Five categories of information relevant to medical research are reported in this manuscript: age, disease/symptom, body structure, medication name, and medication dosage. Across these five categories, the median number of annotations per report was 24.

For Microsoft Text Analytics for Health, Amazon Medical Comprehend, and IQVIA API Marketplace, plain text data from the TAC 2017 test set or medical transcription reports were sent to the API. For GPT-3.5-turbo and GPT-4, the OpenAI API was provided with prompts containing instructions for MedDRA coding alongside plain text source data (Table 1). No additional fine-tuning was performed. APIs were accessed between October 2022 and August 2023.

**Table 1.** Prompts used to generate outputs from GPT-4 and GPT-3.5-turbo for evaluation of medical natural language processing tasks.

| Evaluation task | Prompt |
| --- | --- |
| MedDRA preferred term coding task | Extract all text spans related to MedDRA listed in from input medical text. Output the results into a python list containing all text spans that have been extracted.<br>Example session:<br>`''' User input: "information: * Peripheral Neuropathy [see Warnings and Precautions ( 5.1 ) ] * Anaphylaxis and Infusion Reactions [see Warnings" You output: ['Peripheral Neuropathy', 'Anaphylaxis', 'Infusion Reactions'] '''` |
| Medical entity extraction from medical transcription reports | Forget about our conversation history and treat this as a brand new task.<br>Extract all text spans corresponding to only the entities listed in ["Age", "Gender", "Disease Or Symptom", "Dosage", "Medication Name", "Body Structure"] from input medical text. Output the results into a Python dictionary for all the entities that have been extracted. Keys for the Python dictionary should be only from this list: ["Age", "Gender", "DiseaseOrSymptom", "Dosage", "MedicationName", "BodyStructure"].<br>Example one:<br>`''' User input: "Patient is a 15 years old gentleman with lung cancer. He is now on chemotherapy." You output: {'Age': ['15 years old'], 'Gender': ['gentleman'], 'BodyStructure': ['lung'], 'DiseaseOrSymptom': ['lung cancer']} '''`<br>Example two:<br>`''' User input: "in the day on Augmentin 400 mg twice daily, Lortab or Tylenol p.r.n." You output: {'Dosage': ['400 mg twice daily'], 'MedicationName': ['Augmentin', 'Lortab', 'Tylenol']} '''` |

Abbreviations: GPT, generative pre-trained transformer; MedDRA, Medical Dictionary for Regulatory Activities

Sensitivity, positive predictive value (PPV), and the macro-averaged F1 score (i.e., the harmonic mean of sensitivity and PPV) were calculated based on API outputs. All three metrics range from 0 to 1, with higher scores representing better performance. After observing GPT-3.5-turbo and GPT-4 outputs varied with identical prompting (temperature hyperparameter was set to 0), we repeated experiments five times and reported the range in outcome metrics. For GPT-4, sensitivity analyses were performed to compare several prompting strategies (see Table 1 in the Supplement).

## Results

Table 2 presents results for the MedDRA coding task. According to F1 scores, which combine sensitivity and PPV into a single metric, GPT-4 had similar performance (5-trial range: 0.67 to 0.73) to healthcare NLP comparators (Microsoft Text Analytics for Health: 0.66; IQVIA API marketplace 0.72); however, the sensitivity of GPT-4 tended to be lower and PPV tended to be higher. With F1 scores of 0.50 to 0.51, GPT-3.5-turbo had considerably lower performance than other comparators.

**Table 2.** Algorithm performance on coding free-text adverse drug reactions^1^ to the MedDRA preferred term ontology.

|  | **Sensitivity** | **Positive predictive value** | **F1 score** |
| --- | --- | --- | --- |
| GPT-3.5-turbo<sup>2,3</sup> | 0.43 to 0.45 | 0.57 to 0.59 | 0.50 to 0.51 |
| GPT-4<sup>3</sup> | 0.58 to 0.70 | 0.75 to 0.81 | 0.67 to 0.73 |
| Microsoft Text Analytics for Health | 0.76 | 0.58 | 0.66 |
| IQVIA API Marketplace | 0.78 | 0.66 | 0.72 |

Abbreviations: GPT, generative pre-trained transformer; MedDRA, Medical Dictionary for Regulatory Activities
Note: The Amazon Comprehend Medical algorithm is not able to perform this task.
<sup>1</sup>Task 4 of the 2017 Text Analytics Coding challenge
<sup>2</sup> GPT-3.5-turbo is equivalent to ChatGPT
<sup>3</sup> For GPT-3.5-turbo and GPT-4, we report the range of metrics observed after 5 trials of providing an identical prompt to the model APIs with temperature set to zero.

The performance of tools in extracting specific categories of information from medical transcription reports is presented in Table 3. Performance of the general LLMs varied widely across data categories; the highest F1 scores were for extraction of age (GPT-3.5-turbo: 0.82 to 0.83, GPT-4: 0.84 to 0.87) and medication name (GPT-3.5-turbo: 0.55 to 0.59, GPT-4: 0.70 to 0.76), while F1 scores for disease/symptom, body structure, and medication dosage were lower than those observed for the healthcare NLP tools. For nearly all categories, GPT-4 outperformed GPT-3.5-turbo; though they had similar F1 scores for the medication dosage task (GPT-3.5-turbo: 0.28 to 0.33; GPT-4: 0.26 to 0.31). Compared to the healthcare NLP tools, GPT-3.5-turbo and GPT-4 generally had lower sensitivity, indicating that these models were more likely to miss relevant medical concepts.

**Table 3.** Algorithm performance on extracting five categories of medical entities from de-identified medical transcription reports.

|  | **Sensitivity** | **Positive predictive value** | **F1 score** |
| --- | --- | --- | --- |
|  | ***Age*** |  |  |
| GPT-3.5-turbo<sup>1</sup> | 0.70 to 0.72 | 0.98 to 0.98 | 0.82 to 0.83 |
| GPT-4<sup>1</sup> | 0.75 to 0.79 | 0.96 to 0.96 | 0.84 to 0.87 |
| Microsoft Text Analytics for Health | 0.82 | 0.94 | 0.88 |
| Amazon Comprehend Medical<sup>2</sup> | -- | -- | -- |
| IQVIA API Marketplace | 0.98 | 1.00 | 0.99 |
|  | ***Disease/symptom*** |  |  |
| GPT-3.5-turbo<sup>1</sup> | 0.22 to 0.23 | 0.45 to 0.49 | 0.30 to 0.31 |
| GPT-4<sup>1</sup> | 0.25 to 0.27 | 0.47 to 0.51 | 0.33 to 0.35 |
| Microsoft Text Analytics for Health | 0.62 | 0.37 | 0.46 |
| Amazon Comprehend Medical | 0.55 | 0.41 | 0.47 |
| IQVIA API Marketplace | 0.75 | 0.83 | 0.83 |
|  | ***Body structure*** |  |  |
| GPT-3.5-turbo<sup>1</sup> | 0.15 to 0.23 | 0.50 to 0.54 | 0.23 to 0.32 |
| GPT-4<sup>1</sup> | 0.29 to 0.33 | 0.55 to 0.56 | 0.38 to 0.41 |
| Microsoft Text Analytics for Health | 0.45 | 0.61 | 0.52 |
| Amazon Comprehend Medical | 0.60 | 0.55 | 0.57 |
| IQVIA API Marketplace | 0.96 | 0.86 | 0.91 |
|  | ***Medication name*** |  |  |
| GPT-3.5-turbo<sup>1</sup> | 0.52 to 0.54 | 0.56 to 0.67 | 0.55 to 0.59 |
| GPT-4<sup>1</sup> | 0.68 to 0.73 | 0.73 to 0.78 | 0.70 to 0.76 |
| Microsoft Text Analytics for Health | 0.73 | 0.81 | 0.77 |
| Amazon Comprehend Medical | 0.73 | 0.79 | 0.76 |
| IQVIA API Marketplace | 0.98 | 0.59 | 0.73 |
|  | ***Medication dosage*** |  |  |
| GPT-3.5-turbo<sup>1</sup> | 0.42 to 0.54 | 0.21 to 0.23 | 0.28 to 0.33 |
| GPT-4<sup>1</sup> | 0.38 to 0.46 | 0.19 to 0.24 | 0.26 to 0.31 |
| Microsoft Text Analytics for Health<sup>2</sup> | -- | -- | -- |
| Amazon Comprehend Medical | 0.50 | 0.45 | 0.47 |
| IQVIA API Marketplace | 0.85 | 0.88 | 0.86 |

Abbreviations: GPT, generative pre-trained transformer
<sup>1</sup> For GPT-3.5-turbo and GPT-4, we report the range of metrics observed after 5 trials of providing an identical prompt to the model APIs with temperature set to zero.
<sup>2</sup> The Amazon Comprehend Medical tool is not able to extract age and the Microsoft Text Analytics for Health tool is not able to extract medication dosage.

Experiments comparing GPT-4 prompting strategies showed single sentence examples tended to yield better performance than providing a full medical transcription report (see eTable 2).

## Discussion

Automation of healthcare NLP tasks like information extraction and medical coding require high accuracy, as results can impact patient care, clinical diagnoses, or regulatory decision making. Our results add to an emerging body of evidence that performance of general purpose LLMs varies considerably between models and healthcare tasks.^9^ Despite lack of fine-tuning for medical tasks, GPT-4 performed similarly to healthcare-specific NLP tools on MedDRA coding adverse drug reactions and some medical concept extractions. For other medical concepts, GPT-4 had substantially worse performance than comparators. GPT-3.5-turbo was consistently less accurate than alternatives.

LLMs are trained for use across a broad range of applications but may not be optimal for specialized use cases without domain adaptation. For answering multiple choice and open-ended medical questions, domain fine-tuning a general LLM can lead to substantial performance improvements.^12^ Prompt engineering, or improving model outputs by curating the instruction text for a specific task, can also benefit performance.^13^ The tendency of LLMs to produce plausible but factually incorrect statements, often called ‘hallucination’,^14^ introduces new challenges when considering deployment in healthcare. Differing outputs from GPT-3.5-turbo and GPT-4 models with identical prompts hampers reproducibility and may present practical complications for their use.

As reflected by FDA guidance, it is essential to measure accuracy of new technologies before evaluating their appropriateness for specific tasks.^15^ While this study facilitates side-by-side comparisons of these five algorithms on specific tasks relevant to clinical research, there are several important limitations.

First, this study did not systematically evaluate techniques or strategies to improve LLM performance; however, sensitivity analyses compared several simple prompting strategies. Second, numerous open-source and commercially available alternatives can be used to perform the tasks in this brief report; the three comparators provide a general sense of specialized healthcare NLP performance, results which align with online reports of additional products.^16^ Third, we focused on two NLP tasks pertinent to clinical research; many other tasks could be benchmarked and would provide further understanding of the capabilities of LLMs. Finally, for the medical transcription report extraction task, IQVIA API Marketplace tools were created using similar reports to those used in this evaluation. While the specific examples used in this evaluation were never used for tool development, this may explain IQVIA API Marketplace’s higher performance in Table 3 compared to other healthcare NLP algorithms.

Determining to what extent LLMs can safely and effectively be used to facilitate clinical care, assist in biomedical research, and enable automation of medical administrative burden is still unclear. Making evidence-based decisions requires robust performance evaluation and benchmarking, and these findings contribute to a growing evidence base. When used for NLP tasks commonly performed in clinical research, currently available general LLMs may need fine-tuning on medical data to exceed performance currently observed for healthcare-specific NLP tools.

## Data Availability

All algorithms compared in this manuscript were commercially available at the time of article submission. Data from the 2017 TAC Challenge are publicly available. De-identified medical transcription report data used to create in the medical entity extraction evaluation (without ground truth annotations) are publicly available. Code for evaluation could be made be available upon request to authors.

## Acknowledgements

MedDRA® trademark is registered by the International Council for Harmonisation of Technical Requirements for Pharmaceuticals for Human Use.

